# Prognostic value of NT-proBNP in patients with severe COVID-19

**DOI:** 10.1101/2020.03.07.20031575

**Authors:** Lei Gao, Dan Jiang, Xue-song Wen, Xiao-cheng Cheng, Min Sun, Bin He, Lin-na You, Peng Lei, Wei-xiao Tan, Shu Qin, Guo-qiang Cai, Dong-ying Zhang

## Abstract

The outbreak of coronavirus disease 2019 (COVID-19) caused by severe acute respiratory syndrome coronavirus 2 (SARS-CoV-2) in China has been declared a public health emergency of international concern. The cardiac injury was dominate in the process. However, whether N terminal pro B type natriuretic peptide (NT-proBNP) predicted outcome of COVID-19 patients was unknown. The study initially enrolled 102 patients with severe COVID-19 pneumonia from a continuous sample. After screening out the ineligible cases, 54 patients were analyzed in this study. Results found that patients with higher NT-proBNP (above 88.64 pg/mL) level had more risks of in-hospital death. After adjusting for potential cofounders in separate modes, NT-proBNP presented as an independent risk factor of in-hospital death in patients with severe COVID-19.

## Introduction

The outbreak of coronavirus disease 2019 (COVID-19) caused by severe acute respiratory syndrome coronavirus 2 (SARS-CoV-2) in China has been declared a public health emergency of international concern^1^. Despite lower mortality rate, SARS-CoV-2 has killed more people than SARS and MERS and the number keeps growing^2^. Epidemic studies have been well described clinical features of patients with COVID-19, with the disease severity as an independent predictor of poor outcome^3,4^. Investigating prognostic markers for severe patients are required to supply important information for early therapeutic strategy.

A retrospective, single-center case series of the 138 COVID-19 patients study reported that 7.2% and 16.7% patients had complications of acute cardiac injury and arrhythmia respectively^5^. The percentage of acute cardiac injury and arrhythmia is even higher in severe patients with 22.2% and 44.4% respectively. The severe patients also showed higher creatine kinase-MB (CK-MB) and hypersensitive troponin I (hs-TnI) levels than others^5^. A latest study also reported cardiac injury of COVID-19 was common in severe patients with the peak value of TnI over 40 folds than normal value^6^. However, there is no research concerning whether the heart failure marker, N terminal pro B type natriuretic peptide (NT-proBNP) predicted outcome of COVID-19 patients.

## Methods

### Subjects

The study initially enrolled 102 patients with severe COVID-19 pneumonia from a continuous sample in Hubei General Hospital during the management by national medical team. The study is a retrospective, observational registry with ClinicalTrials.gov identifier NCT04292964. All procedures were followed the instructions of local ethic committee (approval NO. 20200701). All the data were collected using a same protocol by well-trained researchers with a double-blind method. Lacking NT-proBNP results (*n*=45) were excluded. Patients who had stroke (*n*=2) and acute myocardial infarction (*n*=1) were excluded. Other inclusion and exclusion criteria were followed the protocol of the registry (NCT04292964). Finally, 54 patients with COVID-19 were studied in this research.

### Baseline data and follow-up

Demographic data, clinical features and medical history were available and collected according to the patient record system. Data collection of laboratory results were defined using the first-time examination at admission. All the laboratory data were tested in a same laboratory with the same standard. To observe the risks of in-hospital death, patients were followed up from admission to discharge (1 to 15 days). The primary outcome was in-hospital death defined as the case fatality rate. The follow-up data was collected from reviewing medical records by trained researchers using double-blind method.

### Statistical analysis

Data are presented as mean ± SEM, frequency (%) or median (interquartile ranges). Intergroup comparisons between NT-proBNP higher group and lower group were made by the independent-samples *T*-test (normally distributed continuous variables), Mann-Whitney *U* test (nonnormally distributed continuous variables) and chi-square test (categorical variables). The best NT-proBNP cut-off was that of the highest product of sensitivity and specificity for in-hospital death prediction. Cumulative survival curves of in-hospital death were estimated using the Kaplan-Meier product-limit estimation method with the log-rank test. Spearman correlation analyzing was used to investigate the coefficients of NT-proBNP with selected covariates. Cox proportional hazards models were used to screening out the potential risk factors and analyzing the independent effect of NT-proBNP on in-hospital death. Statistical analyses were performed by SPSS 22.0 (SPSS, Chicago, IL, USA) and a two-sided *P*<0.05 was considered statistically significant.

## Results

### Baseline characteristics

Baseline characteristics of participants were divided into two groups by lower and higher NT-proBNP levels (NT-proBNP≤88.64 pg/mL and NT-proBNP>88.64 pg/mL, Table 1) according to the cut-off determined in the ROC curve (Figure 1). Patients in NT-proBNP higher group were significantly older with more hypertension (HP) and coronary heart disease (CHD) histories, higher levels of diastolic blood pressure (DBP), myohemoglobin (MYO), CK-MB, hs-TnI, blood urea, creatinine, white blood cell (WBC), C-Reactive Protein (CRP) and procalcitonin (PCT), lower level of lymphocyte (LYM) and higher rate of in-hospital death. Other characteristics like male/female, temperature, pulse rate, respiratory rate, systolic blood pressure, chronic obstructive pulmonary disease (COPD) history and diabetes history showed no insignificance between groups by NT-proBNP (Table 1.).

**Table 1.** Baseline characteristics of total and different degrees of NT-proBNP.

| Characteristics | Total (n=54) | NT-proBNP≤88.64<br>pg/ml (n=24) | NT-proBNP>88.64<br>pg/ml (n=30) | *P* |
| --- | --- | --- | --- | --- |
| Male/Female (n) | 24/30 | 8/16 | 16/14 | 0.142 |
| Age (years) | 60.4±16.1 | 51.6±13.9 | 67.4±14.4 | <0.001 |
| Temperature (°C) | 36.7 (36.5-36.9) | 36.8 (36.5-36.9) | 36.6 (36.5-36.9) | 0.670 |
| Pulse (/min) | 82 (76-97) | 84 (76-97) | 82 (76-96) | 0.679 |
| Respire (/min) | 20 (19-21) | 20 (19-20) | 20 (18-26) | 0.209 |
| SBP (mmHg) | 128 (119-138) | 126 (115-134) | 129 (120-144) | 0.218 |
| DBP (mmHg) | 78 (70-83) | 73 (69-78) | 80 (70-86) | 0.040 |
| History of HP (n) | 12 (22.2%) | 2 (8.3%) | 10 (33.3%) | 0.028 |
| History of CHD (n) | 9 (16.7%) | 1 (4.2%) | 8 (26.7%) | 0.027 |
| History of COPD (n) | 2 (3.7%) | 0 | 2 (6.7%) | 0.197 |
| History of DM (n) | 8 (14.8%) | 3 (12.5%) | 5 (16.7%) | 0.668 |
| NT-proBNP (pg/ml) | 137.30 (39.64-494.98) | 37.28 (22.28-61.74) | 420.40 (199.63-919.88) | <0.001 |
| MYO (ng/ml) | 39.28 (26.26-86.84) | 25.35 (14.04-35.20) | 82.53 (34.55-123.96) | <0.001 |
| CK-MB (ug/L) | 1.04 (0.65-2.27) | 0.63 (0.37-0.79) | 1.90 (1.08-3.78) | <0.001 |
| Hs-TnI (ng/ml) | <0.006 (<0.006-0.022) | <0.006 (<0.006-<0.006) | 0.021 (<0.006-0.136) | 0.001 |
| Urea (mmol/L) | 4.8 (3.3-9.0) | 3.4 (2.6-4.9) | 7.1 (4.4-9.9) | <0.001 |
| Creatinine (umol/L) | 63 (44-77) | 54 (41-69) | 79 (55-86) | 0.016 |
| WBC (10<sup>9</sup>/L) | 5.42 (4.13-7.45) | 5.89 (4.53-10.76) | 5.73 (4.50-8.20) | 0.007 |
| LYM (10<sup>9</sup>/L) | 1.12±0.52 | 1.30±0.43 | 0.98±0.55 | 0.021 |
| CRP (mg/L) | 34.8 (5.3-61.0) | 7.6 (5.0-34.8) | 54.3 (14.3-117.9) | 0.003 |
| PCT (ng/ml) | 0.063 (0.029-0.171) | 0.038 (0.020-0.058) | 0.137 (0.049-0.468) | <0.001 |
| In-hospital death (n) | 18 (33.3%) | 0 | 18 (60.0%) | <0.001 |

Abbreviations: SBP, systolic blood pressure; DBP, diastolic blood pressure; HP, hypertension; CHD, coronary heart disease; COPD, chronic obstructive pulmonary disease; DM, diabetes mellitus; NT-proBNP, N-terminal pro-brain natriuretic peptide; MYO, myoglobin; CK-MB, creatine kinase-MB; Hs-TnI, high-sensitivity troponin-I; WBC, white blood cell; LYM, lymphocytes; CRP, C-reactive protein; PCT, procalcitonin.

**Figure 1.**
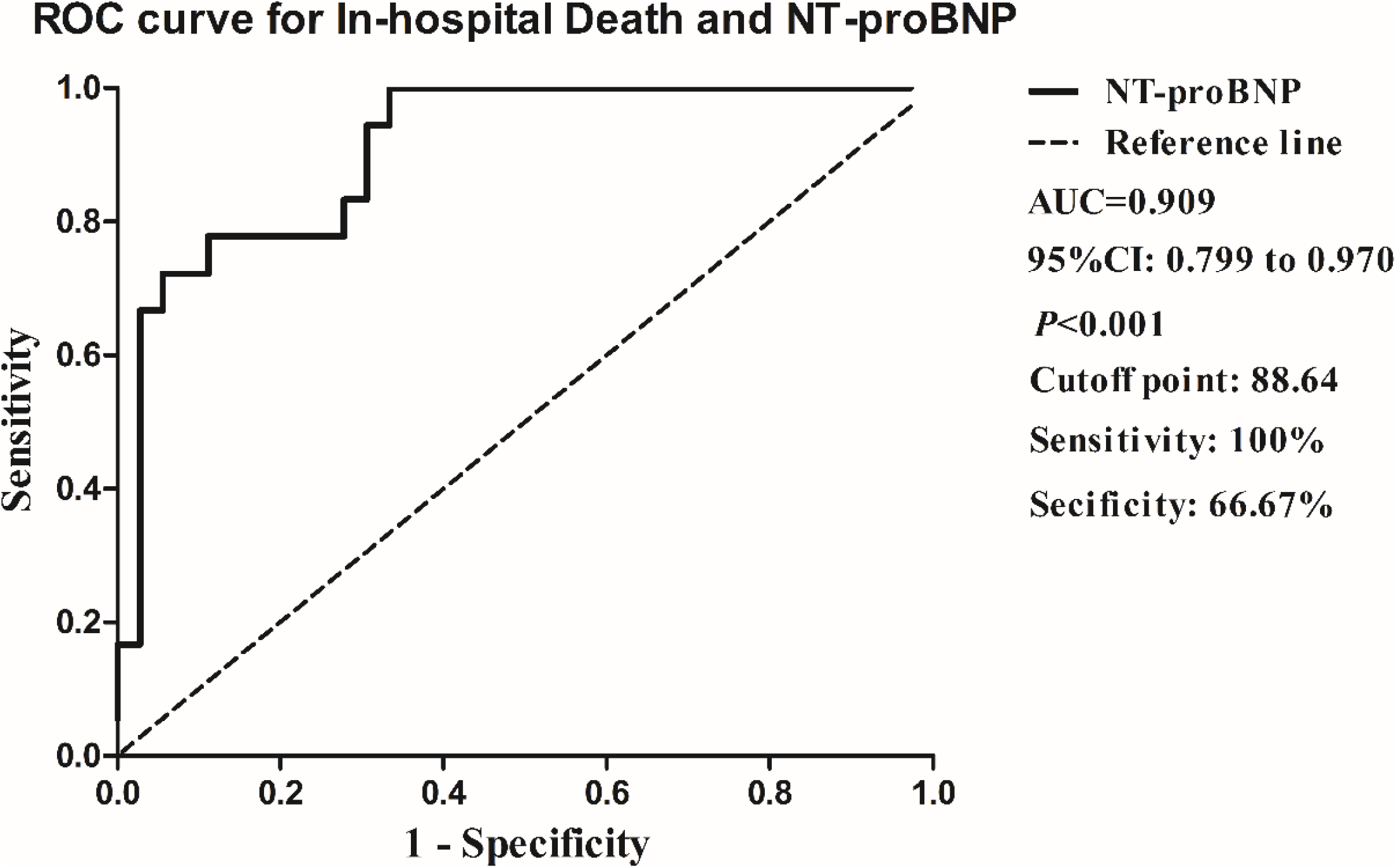
The NT-proBNP for in-hospital death of coronavirus disease 2019 (COVID-19) by receiver operating characteristic (ROC) curves. The area under the curve (AUC) of NT-proBNP was 0.909. The best cutoff of NT-proBNP for prediction in-hospital death was 88.64 pg/mL with the sensitivity of 100% and the specificity of 66.67%. 95%CI, 95% confidence interval.

### Receiver operator characteristic curve (ROC) for prediction in-hospital death

Receiver operation characteristic (ROC) curves were shown in figure 1 to analyze the prognostic value and the best cutoff of NT-proBNP for prediction in-hospital death. The area under the curve (AUC) for in-hospital death was 0.909 (95%CI 0.799-0.970, *P*<0.001). The best cutoff of NT-proBNP for prediction in-hospital death was 88.64 pg/mL with the sensitivity of 100% and the specificity of 66.67% (Figure 1).

### Cumulative survival curves of in-hospital death

Cumulative survival rate curves between groups categorized by NT-proBNP cutoff were shown in figure 2. Patients in higher NT-proBNP (≥88.64 pg/mL) group had a significant higher risk of death during the days of following-up than the other group (NT-proBNP<88.64 pg/mL) (Figure 2).

**Figure 2.**
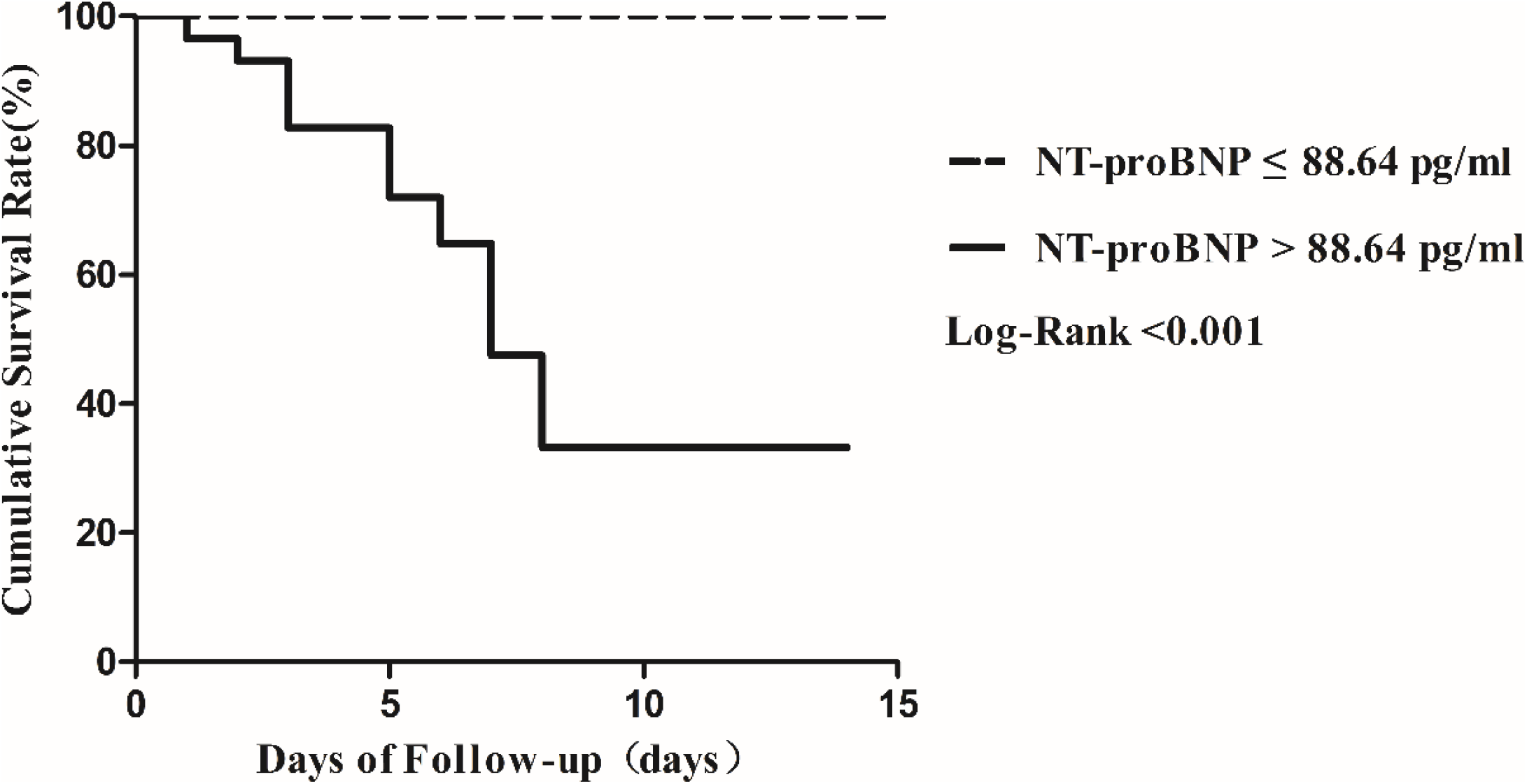
Kaplan-Meier plots showing the cumulative survival rate of COVID-19 patients who were stratified into two groups according to plasma NT-proBNP cutoff point at baseline. (Dotted line, NT-proBNP ≤88.64 pg/ml, n=24; Solid line, NT-proBNP >88.64 pg/ml, n=30; log-rank test for trend, *P*<0.001).

### Spearman correlation coefficients of NT-proBNP with selected covariates

In present study, plasma NT-proBNP was positively correlated with age, Ur, cardiac injury markers of MYO, CK-MB and hs-TnI and systematic inflammation makers of WBC, CRP, Hs-CRP and PCT (Supplemental figure 1).

### Results of Cox proportional hazards analyses of in-hospital death

Cox proportional hazards regression analysis was used to evaluate potential associations between NT-proBNP and in-hospital death. Results of univariate analyses showed that the HR associated to in-hospital death of NT-proBNP was 1.368 (95% CI 1.217-1.541, P<0.001). Meanwhile, older, male, history of hypertension (HP), myoglobin (MYO), creatine kinase-MB (CK-MB), high-sensitivity troponin-I (Hs-TnI), white blood cell (WBC), lymphocytes (LYM), c-reactive protein (CRP) and procalcitonin (PCT) were correlated with the risk of in-hospital death (Table 2)

**Table 2.** Results of univariate Cox proportional-hazards regression analyzing the effect of baseline variables on in-hospital death.

| Characteristics |  | HR (95%CI) | *P* |
| --- | --- | --- | --- |
| Sex | Male | - |  |
|  | Female | 0.348 (0.130-0.930) | 0.035 |
| Age, per 10 years |  | 1.975 (1.309-2.981) | 0.001 |
| History of HP | no | - | - |
|  | yes | 4.044 (1.604-10.200) | 0.003 |
| History of CHD | no | - | - |
|  | yes | 2.652 (0.992-7.092) | 0.052 |
| History of COPD | no | - | - |
|  | yes | 4.127 (0.945-18.024) | 0.059 |
| History of DM | no | - | - |
|  | yes | 0.958 (0.277-3.314) | 0.947 |
| NT-proBNP, per 100 pg/ml |  | 1.369 (1.217-1.541) | <0.001 |
| MYO, per 1 ng/ml |  | 1.006 (1.003-1.008) | <0.001 |
| CK-MB, per 1 ug/L |  | 1.259 (1.098-1.443) | 0.001 |
| Hs-TnI, per 1 ng/ml |  | 1.862 (1.273-2.722) | 0.001 |
| Urea, per 1 mmol/L |  | 1.134 (1.073-1.198) | <0.001 |
| Creatinine, per 1 umol/L |  | 1.028 (1.013-1.043) | <0.001 |
| WBC, per 1x10<sup>9</sup>/L |  | 1.150 (1.076-1.229) | <0.001 |
| LYM, per 1x 10<sup>9</sup>/L |  | 0.065 (0.017-0.249) | <0.001 |
| CRP, per 1 mg/L |  | 1.021 (1.012-1.030) | <0.001 |
| PCT, per 0.1 ng/ml |  | 1.241 (1.142-1.349) | <0.001 |

Abbreviations: HP, hypertension; CHD, coronary heart disease; COPD, chronic obstructive pulmonary disease; DM, diabetes mellitus; NT-proBNP, N-terminal pro-brain natriuretic peptide; MYO, myoglobin; CK-MB, creatine kinase-MB; Hs-TnI, high-sensitivity troponin-I; WBC, white blood cell; LYM, lymphocytes; CRP, c-reactive protein; PCT, procalcitonin; HR, hazards ratio; 95%CI, 95% confidence interval.

Modes of Cox proportional hazards regression analyses were used to evaluate the independent prognostic effect of NT-proBNP level. After adjusting for sex and age (Mode 1), the HR of NT-proBNP for in-hospital death was 1.323 (95% CI 1.119-1.563, P=0.001). After adjusting for HP and CHD history (Mode 2), the HR was 1.342 (95% CI 1.185-1.520, P<0.001). After adjusting for MYO, CK-MB, and hs-TNI (Mode 3), the HR was 1.360 (95% CI 1.177-1.572, P<0.001). After adjusting for urea and creatinine (Mode 4), the HR was 1.373 (95% CI 1.188-1.586, P<0.001). After adjusting for WBC and LYM (Mode 5), the HR was 1.248 (95% CI 1.097-1.419, P=0.001). After adjusting for WBC, LYM and CRP (Mode 6), the HR was 1.230 (95% CI 1.003-1.509, P=0.047). After adjusting for WBC, LYM and PCT (Mode 7), the HR was 1.200 (95% CI 1.045-1.380, P=0.010). In this process, the HRs of WBC and PCT in Mode 5 and 7 also showed significance for independently predicting in-hospital death while LYM show protective effect (Table 3, Figure 3).

**Table 3.**
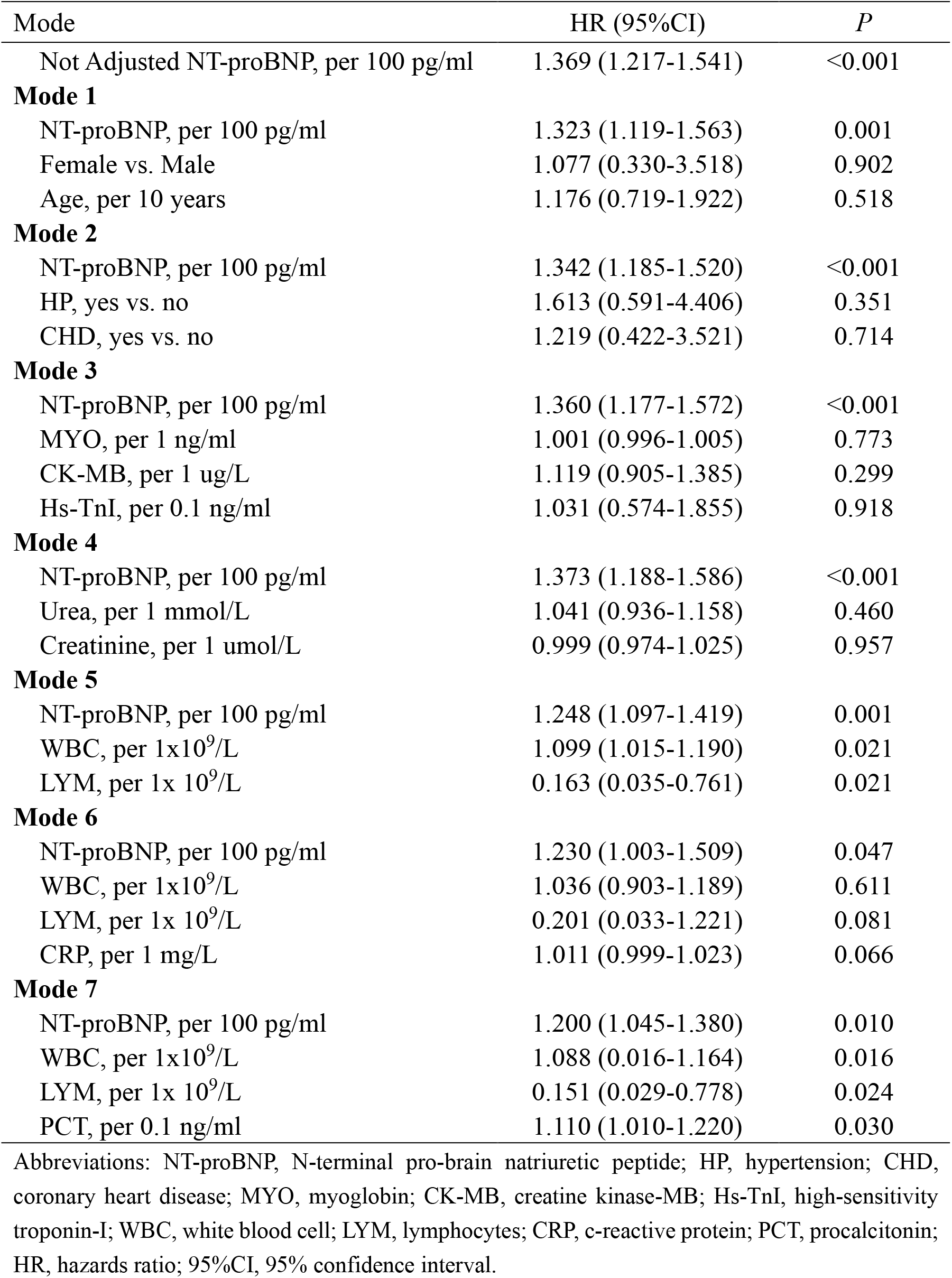
Results of multivariate Cox proportional-hazards regression analyzing the effect of baseline variables on in-hospital death.

**Figure 3.**
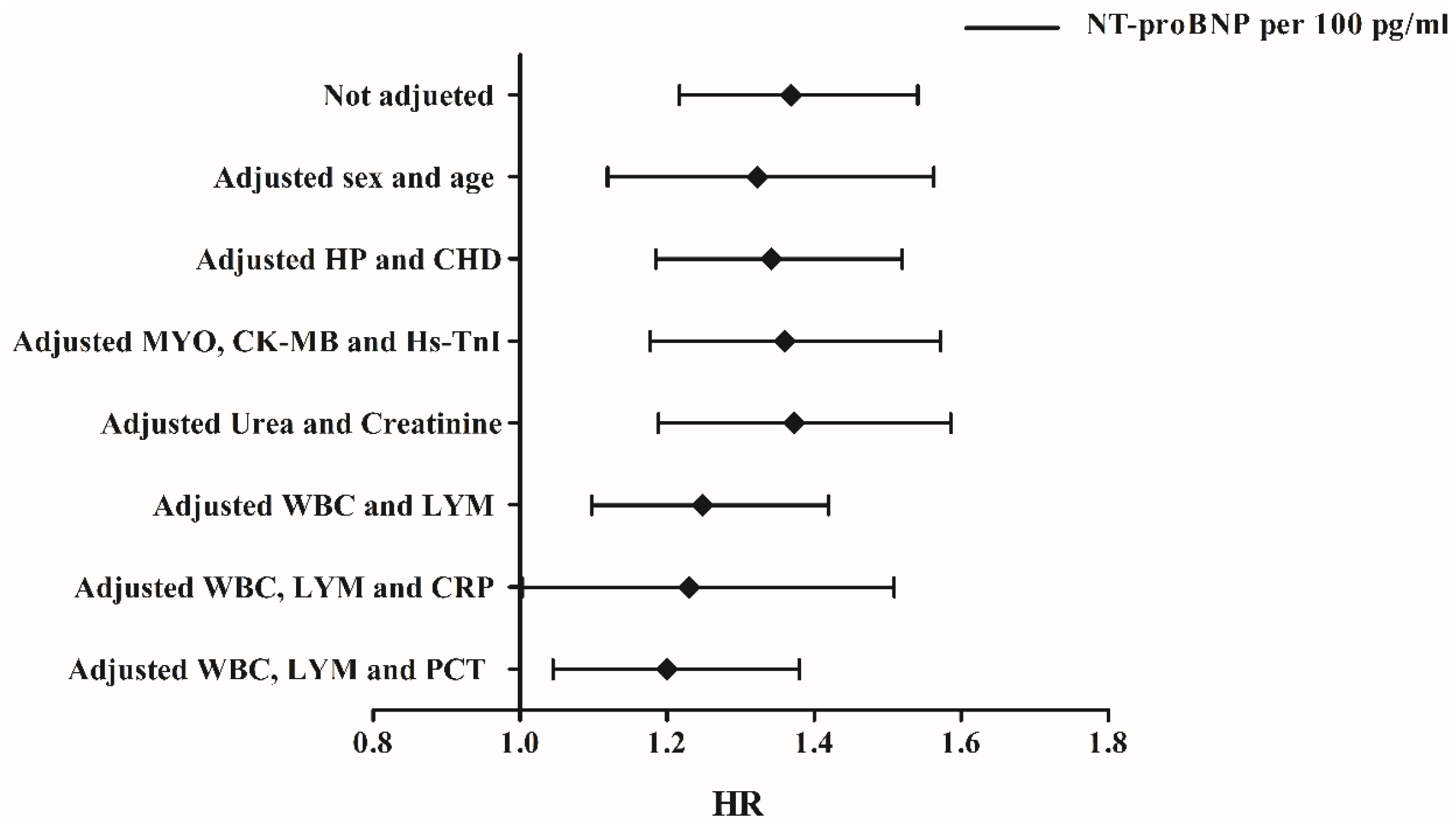
Forest plots of multivariate Cox proportional-hazards regression analyzing the effect of baseline variables on in-hospital death. HP, hypertension; CHD, coronary heart disease; MYO, myoglobin; CK-MB, creatine kinase-MB; Hs-TnI, high-sensitivity troponin-I; WBC, white blood cell; LYM, lymphocytes; CRP, c-reactive protein; PCT, procalcitonin; HR, hazards ratio.

## Discussion

The present study for the first time showed the relationship between plasma NT-proBNP level and risks of in-hospital death in severe COVID-19 patients. Severe COVID-19 patients with higher NT-proBNP levels tended to be older with higher levels of cardiac injury markers and higher levels of systematic inflammation markers. Patients with higher NT-proBNP (above 88.64 pg/mL) level had lower cumulative survival rate. After adjusting for potential cofounders in separate modes, NT-proBNP presented as an independent risk factor of in-hospital death in patients with severe COVID-19.

Previous studies have found that NT-proBNP is a powerful and independent predictor of mortality in community-acquired pneumonia (CAP)^7-9^. In these studies the best cutoffs of NT-proBNP for prediction 30-day mortality were 1,434.5 pg/mL and 1,795.5 pg/mL respectively^7,9^. The elevated NT-proBNP in these cases was believed owing to the cardiac complications resulted from complex interactions among preexisting conditions, relative ischemia, up-regulation of the sympathetic system, systemic inflammation and direct pathogen-mediated damage to the cardiovascular system^10^.

However, in present study, the cutoff value of NT-proBNP to predict the adverse outcome of severe COVID-19 patients was far lower than the threshold to diagnose heart failure (450 pg/mL for <50 years, 900 pg/mL for 50-75 years and 1,800 pg/mL for >75 years)^11^. It was suggested that the prognostic effect of plasma NT-proBNP in severe COVID-19 patients could not fully ascribe to heart failure induced by the virus or hypoxia. Further understanding of physiological and pathological significance of plasma NT-proBNP elevation in severe COVID-19 patients might help clinicians make corresponding decisions to reduce the risks of adverse outcome.

The prognostic effect of NT-proBNP might indicate the extent of cardiac stress and inflammation. Previous reports have pointed that heart injury of severe patients with new coronary pneumonia is very prominent and cardiac death has become an important cause of death of patients with COVID-19^5,6^. The mechanism of COVID-19 associated cardiac injury was still unclear. From the result of autopsy by Xu and colleagues, a few interstitial mononuclear inflammatory infiltrates were observed in heart biopsy, indicating an inflammation induced cardiac injury^12^. Other factors including the SARS-CoV-2 infection and invasion cardiomyocytes via the binding site of angiotensin-converting enzyme-related carboxypeptidase (ACE2)^13^, the pulmonary infection induced inadequate oxygen supply to the myocardium and the influences of cytokine storm syndrome ^14-16^ might also contribute to the cardiac injury^17^.

In present study, NT-proBNP was positively correlated to the makers of cardiac injury and systematic inflammation. The levels of cardiac injury and inflammation also showed risks of in-hospital death by univariate Cox proportional-hazards regression analyze. However, in multiple regression analyses, the cardiac injury markers were not associated with the hazard ratios of in-hospital death under the affection of NT-proBNP. Both NT-proBNP and inflammation showed independent effect of predicting in-hospital death. Considering these, the prognostic effect of NT-proBNP might be a comprehensive index of reflecting the overall state of the organism.

SARS-CoV-2 binds with ACE2, resulting the uncontrolled releasing of angiotensin 2 (ANGII) and restricted synthesis of ANG1-7^18^. The latter exerts anti-inflammation effect to protect tissue while ANGII plays in an opposite role and facilitating the secretion of NT-proBNP^18-20^, indicating the NT-proBNP level might associated with the severity of infection, which needs further verification.

Font-line doctors pointed that some non-survivors with severe COVID-19 pneumonia died without warning. By investigating the prognostic effect of NP-proBNP level of severe COVID-19 patients at admission, it might be helpful to identifying patients with poor prognoses early.

## Data Availability

The raw data required to reproduce these findings cannot be shared at this time as the data also forms part of an ongoing study.

## Acknowledgements

We thank all the staffs working at the front-line to battle against SARS-CoV-2. We show our respect to those who have sacrificed in this war.

